# Varying Malaria Rapid Diagnostic Test Accuracy by Regional Transmission Level and Demographics in Tanzania

**DOI:** 10.1101/2025.07.17.25331734

**Authors:** Danielle Wiener, Misago D. Seth, Celine I. Mandara, Rashid A. Madebe, Zachary R. Popkin-Hall, David Giesbrecht, Catherine Bakari, Beatus Lyimo, Dativa Pereus, Filbert Francis, Daniel Mbwambo, Sijenunu Aaron, Abdalah Lusasi, Samwel Lazaro, Timothy P. Sheahan, Jonathan B. Parr, Jeffrey A Bailey, Deus S. Ishengoma, Jonathan J. Juliano

## Abstract

Malaria remains a significant global health burden, with approximately 263 million cases across 83 countries. It’s essential for malarial infection control to quickly and accurately detect cases. Given the widespread use of malaria rapid diagnostic testing (mRDTs) for case management and surveillance, it’s essential to understand test reliability. Clarifying how mRDT results differ from qPCR results, and the nature of additional variance by test manufacturer, will be useful for reducing measurement bias. In comparing 3 national standard mRDTs and a research mRDT with qPCR results from a 2021 cross-sectional study in Tanzania, differences were found by age, gender and regional malaria transmission rate. The research test overall underperformed, with poor sensitivity across transmission strata. In comparing the research mRDT to standard mRDTs, odds ratios suggested transmission intensity may affect mRDT agreement and diagnostic performance. These results offer pertinent information on test accuracy and decrease outcome misclassification for malaria prevalence.

---

Malaria remains a major public health challenge, affecting approximately 263 million people in 2023. ^1^ Despite long-standing control efforts, transmission persists. A key strategy in malaria control is test-and-treat, which depends on accurate diagnosis and effective treatment. In Africa, malaria rapid diagnostic tests (mRDTs) are the most widely used diagnostic tool due to their speed, affordability, and ease of use.^1^ Microscopy is less common due to technical and cost barriers, while nucleic acid tests like qPCR offer the highest sensitivity but are impractical for routine use. ^2^ However, mRDTs face limitations, including inadequate sensitivity leading to false negatives, false positives due to lingering histidine rich protein 2 (HRP2) antigen after treatment, and reduced sensitivity to non-falciparum infections.^2, 3^ Additionally, *Pfhrp2* and *Pfhrp3* gene deletions in *P. falciparum* have raised concerns about HRP2/3-based mRDT accuracy, as these deletions can result in undetected infections. ^4^

Understanding how mRDTs perform in various settings is essential for their effective implementation. Prior studies have shown that in regions such as Southwest Nigeria and low-transmission areas of Tanzania, the specificity and positive predictive value of mRDTs decline among children under five, often resulting in overdiagnosis. ^2, 5^ Commonly used HRP2/pan-Plasmodium lactate dehydrogenase (pLDH) mRDTs are widely adopted in malaria-endemic countries and generally demonstrate similar diagnostic accuracy. ^6^ However, when compared to more sensitive reference methods like quantitative PCR (qPCR), these tests tend to show reduced specificity and more variable performance, sometimes falling short of World Health Organization (WHO) standards. ^3, 7, 8^

A newer diagnostic tool, the Rapigen Biocredit ™ Malaria Ag Pf, utilizes a PfLDH-based approach and has shown promise in detecting malaria cases, particularly in areas where *Pfhrp2* gene deletions are prevalent. Studies suggest that PfLDH based mRDTs, such as Rapigen may offer improved sensitivity and specificity in regions where hrp2/3 deletions are common, while maintaining comparable accuracy in non-deletion cases. ^9, 10, 11^ However, PfLDH-based mRDTs have also been associated with poor sensitivity and specificity, as well as reduced effectiveness in detecting infections with low parasitemia. ^12, 13^ These limitations highlight the need for further research to evaluate the diagnostic accuracy of the Rapigen and other PfLDH mRDTs relative to standard HRP2-based tests, especially considering variables such as regional transmission intensity and patient age.

This study is a secondary analysis of 3,284 dried blood spot (DBS) samples collected in 2021 through the Molecular Surveillance of Malaria in Mainland Tanzania (MSMT) project. The original study aimed to investigate *Pfhrp2/3* gene deletions, population genetics of Plasmodium parasites and the profile of antimalarial drug resistance markers across ten regions of Tanzania, representing the country’s government-defined malaria transmission strata.^14, 15, 16^ Participants presenting with malaria-like symptoms at health facilities were tested using one or more mRDTs, and DBS samples were collected for molecular analysis. DNA was extracted from the DBS using Chelex extraction and *Plasmodium falciparum* was detected using an assay targeting the 18S ribosomal subunit, as previously described.^17^ Additional methodological details are available elsewhere. ^17, 18^ Individuals who did not receive both the standard and research mRDTs were excluded from this analysis (final sample: n=3,199; see **Table 1**). Standard mRDTs included SD Bioline Malaria Ag P.f/pan (#05FK60, Standard Diagnostic Inc., India), CareStart Malaria HRP2/pLDH (#RMOM-02571, AccessBio Inc., USA), and First Response Malaria Ag HRP2/pLDH Combo (#PI16FRC10s, Premier Medical Corp., India). The research mRDT evaluated was the BIOCREDIT Malaria Ag Pf (Pf-pLDH) (#C14RHG25, RapiGEN Inc., Republic of Korea).

**Table 1.**
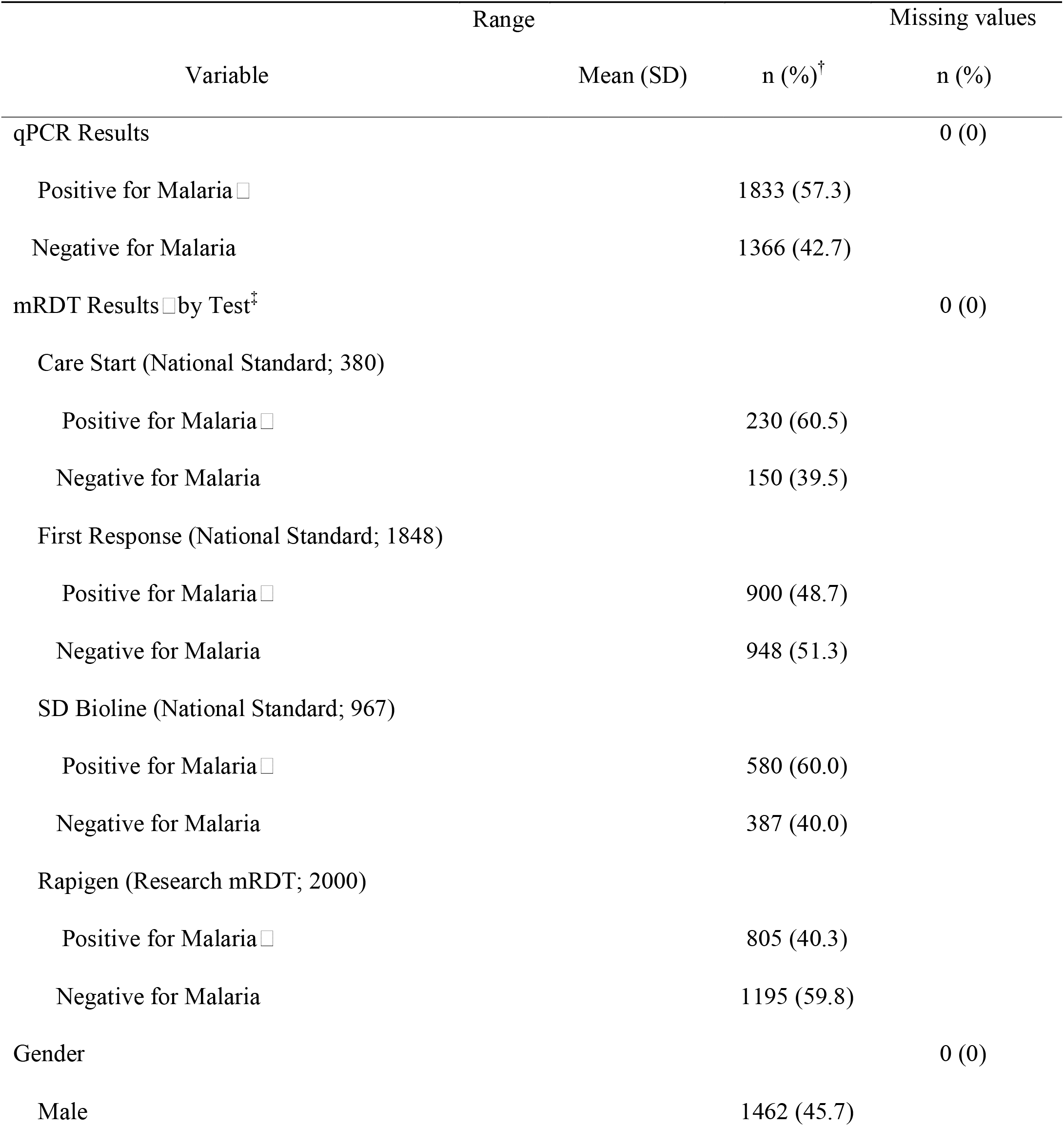
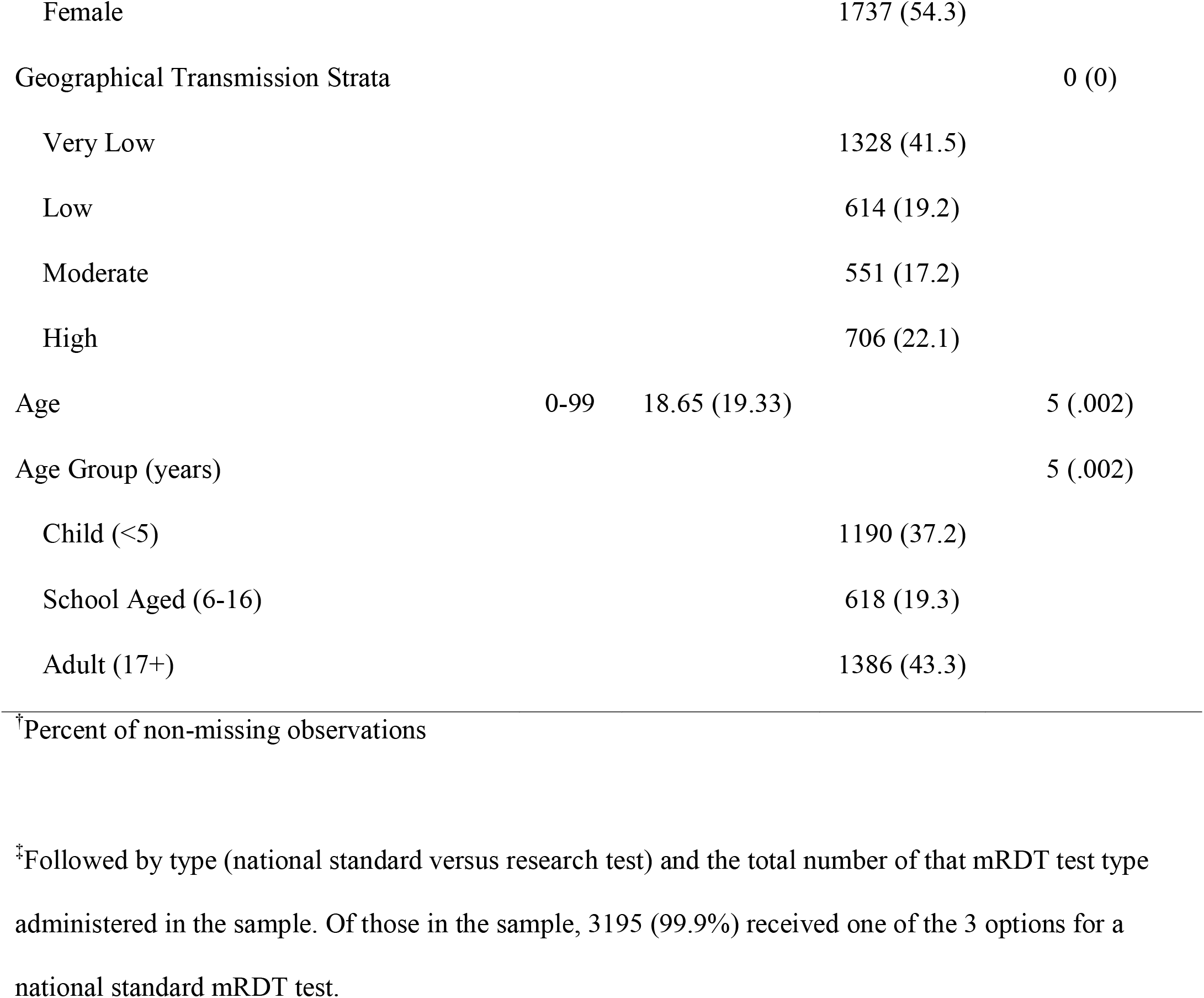
Descriptive Characteristics of Study Sample, Tanzania 2021 (N=3199)

Test accuracy was assessed using sensitivity, specificity, positive predictive value (PPV), and negative predictive value (NPV). Since PPV and NPV depend on disease prevalence, they are not generalizable across settings with different malaria rates. A supplementary analysis applied a parasitemia threshold to qPCR results, treating samples with ≤50 copies as negative, reflecting the typical detection limit of mRDTs and expert microscopy. Results from this threshold-based analysis are presented in Supplemental Tables 1 and 2, alongside unadjusted results. Logistic regression using SAS 9.4 was used to calculate diagnostic odds ratios (ORs) for agreement between the research and standard mRDTs, stratified by transmission intensity (high, moderate, low, very low) based on Tanzania’s 2020 malaria stratification data. ^14^

When stratified by age and gender, test accuracy varied by manufacturer (**Table 2**). The research mRDT (Rapigen PfLDH) showed the lowest sensitivity across all age groups, performing worst in each stratum. Among children under five, CareStart had the highest sensitivity (86.4%), while Rapigen had the lowest (69.7%). Specificity was highest among adults aged 17 and older (91.4–97.9%) across all tests and remained consistently high for the research mRDT across age and sex. PPV was generally high across all strata, with First Response being the only test to consistently fall below 0.900. In contrast, NPV was the weakest metric overall, with 13 of 20 estimates showing NPV below 0.800. The research mRDT had the lowest NPV in nearly every group. Overall, demographic stratification revealed that while the research mRDT had relatively high specificity, it consistently underperformed in sensitivity.

**Table 2.**
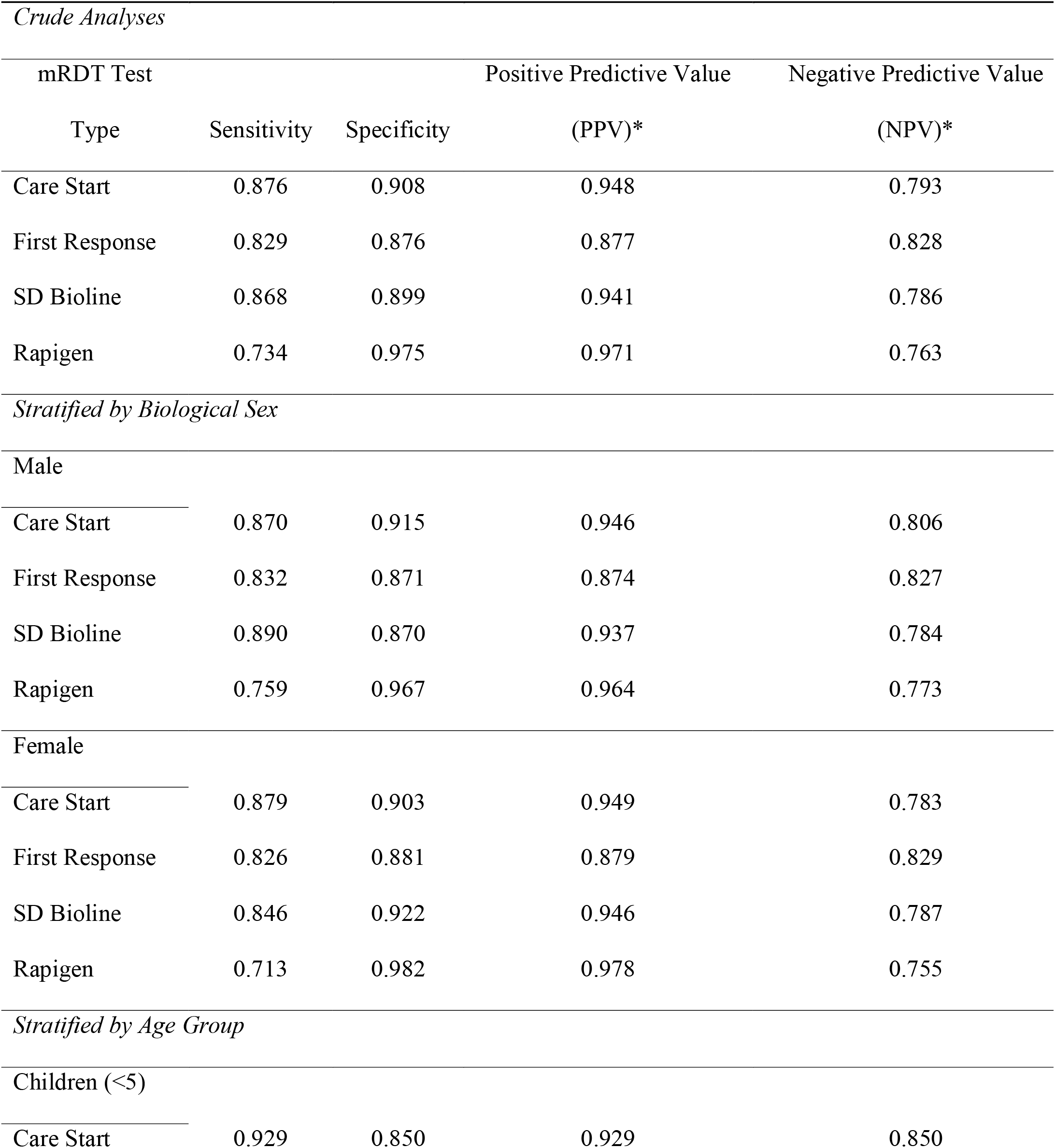
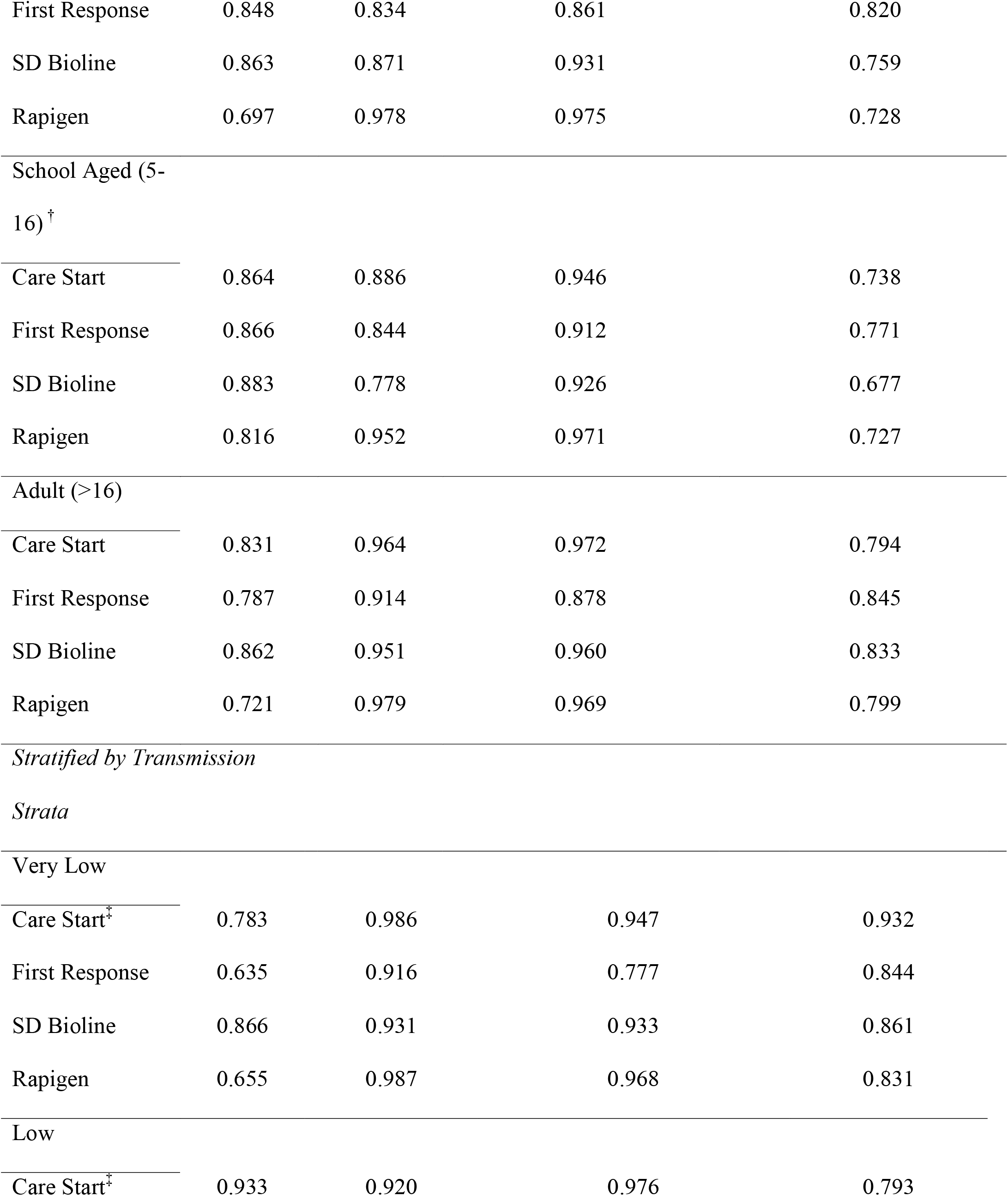
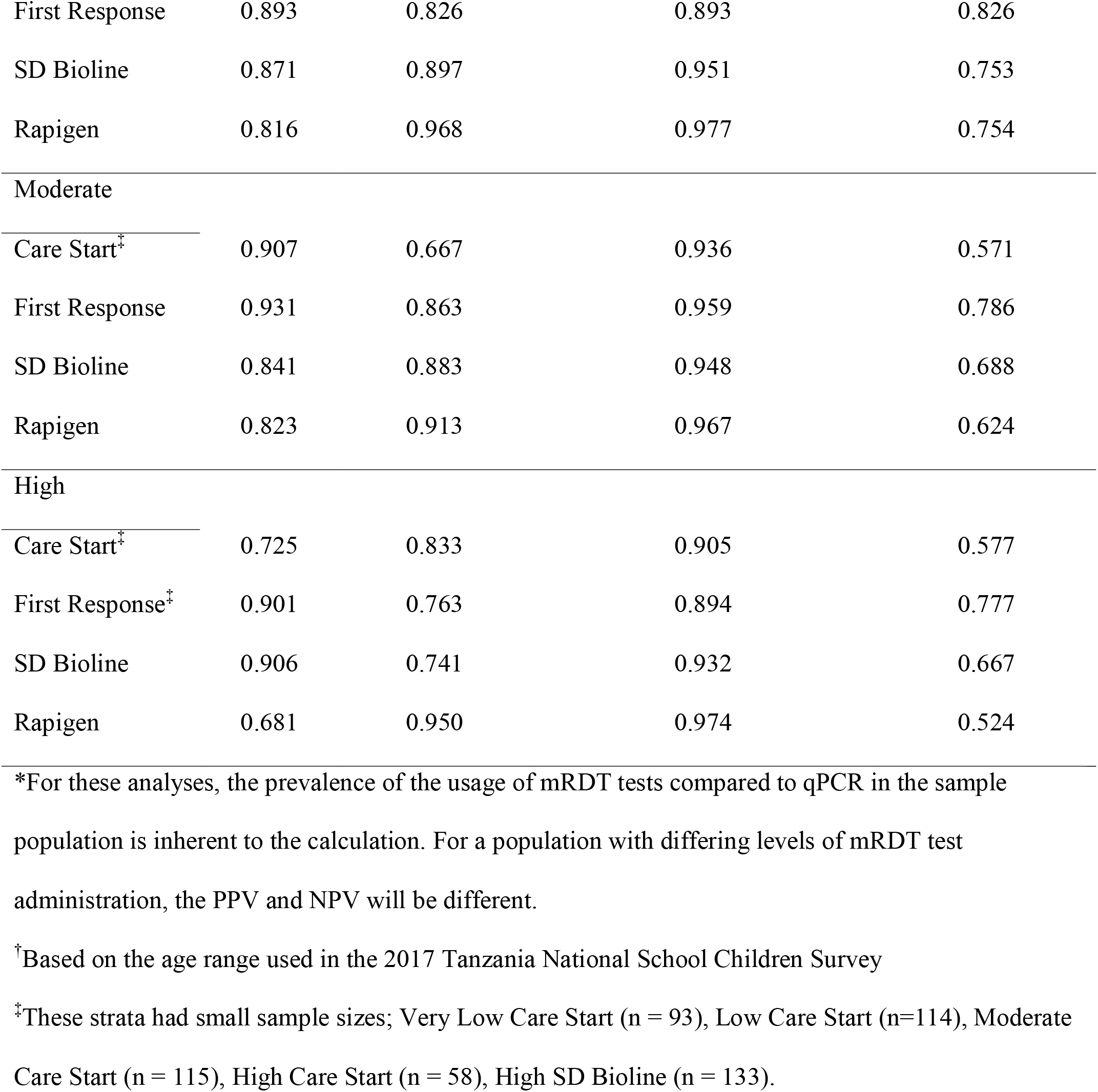
Accuracy of mRDTs Compared to qPCR by Sex, Age Group and Transmission Strata.

Regional malaria transmission levels appeared to influence mRDT accuracy, with variation by test manufacturer. CareStart and First Response showed higher sensitivity in low to moderate transmission areas, while First Response performed poorly in very low transmission regions (sensitivity = 63.5%, PPV = 77.7%). SD Bioline had reduced specificity in high transmission areas (74.1%). PPV remained high across all strata and tests, while NPV was generally low except in very low transmission areas. Although diagnostic odds ratios (ORs) for agreement between the research and standard mRDTs varied by region, overlapping confidence intervals suggest limited statistical difference (**Figure 1**). Overall, the odds of a positive malaria diagnosis from the standard mRDTs among those who were diagnosed positive from the PfLDH-based test were 216.8 (95% CI: 138.7, 338.8) times the odds of those who were diagnosed negative from the PfLDH-based test – indicating high sensitivity and agreement. In very low to moderate regions, the odds of a positive result on a standard test given a positive research test ranged from 203.4 to 244.4 (95% CI: 62.7–740.8). The high transmission region showed a lower OR estimate (84.4; 95% CI: 29.4–242.1), suggesting that transmission intensity may affect mRDT agreement and diagnostic performance.

**Figure 1.**
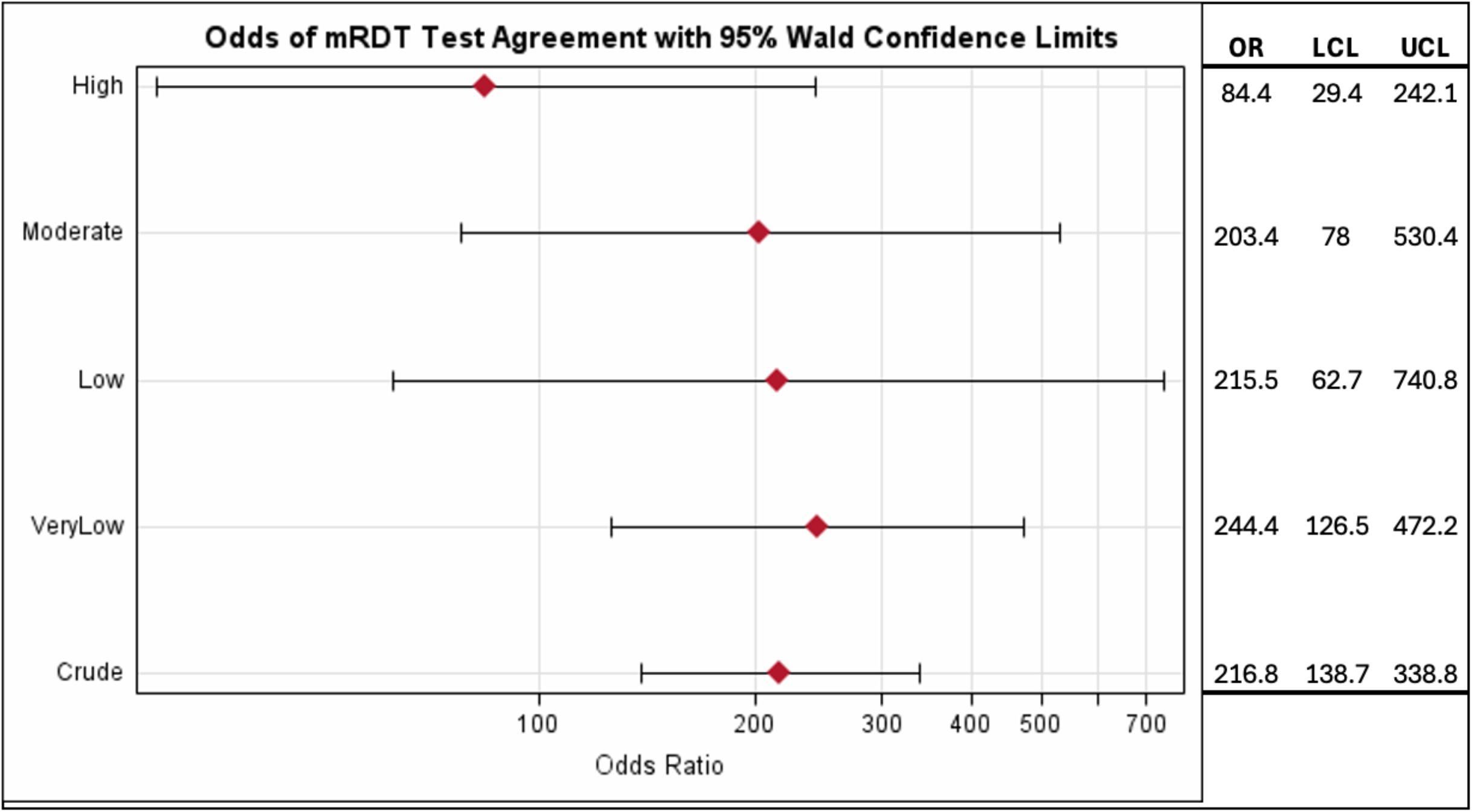
Odds Ratio for Agreement Between Rapigen Test and Standard mRDTs. The likelihood of agreement between the research Rapidgen test and standard mRDT are shown stratified by transmission intensity. OR: Odds Ratio; LCL: Lower Confidence Interval; UCL: Upper Confidence Interval. All ORs have a p-value <.0001.

Applying a qPCR parasitemia threshold of ≥50 copies (**Supplemental Tables 1 and 2**) did not meaningfully alter results, with accuracy estimates changing by no more than 2 percentage points—except for the CareStart mRDT, which had a smaller sample size (n = 380). This limited sample may reduce the reliability of its accuracy estimates.

Key limitations of this study include the small sample size for CareStart and wide confidence intervals from logistic regression, indicating lower precision. A larger sample would likely improve estimate stability, as seen in the overall crude odds ratio (**Figure 1**) compared to stratified analyses. Additionally, all samples were collected from symptomatic individuals, limiting generalizability to asymptomatic or non-health-seeking populations. However, mRDTs are meant to be used for the diagnosis of symptomatic malaria.

This study highlights notable differences in diagnostic performance between standard mRDTs and the newer Rapigen test. While Rapigen showed high specificity—potentially reducing overtreatment—it consistently underperformed in sensitivity and negative predictive value, especially in high transmission areas. These limitations are concerning, as missed infections can lead to untreated cases and sustained transmission. Accurate diagnosis is essential for malaria control, particularly as *Pfhrp2/3* gene deletions and rising drug resistance challenge current strategies. While sensitivity ensures timely treatment, specificity helps avoid unnecessary drug use and resistance. A reliable diagnostic must perform well across diverse populations and settings. These findings underscore the need for ongoing evaluation of mRDTs as malaria transmission patterns and parasite genetics evolve. Strengthening diagnostic tools is critical to support test-and-treat programs and advance toward malaria elimination goals.

**Alt Text Table 1** = Table showing demographic characteristics of the Tanzania study population, including qPCR Results, mRDT Results by manufacturer, gender, transmission strata and age group. There are no missing values except for .002% in the age variable. The majority of the study population lives in very low transmission strata (42%), are female (54%), >16 years old (43%) and qPCR positive for malaria (57%).

**Alt Text Table 2** = Table giving estimates of sensitivity, specificity, PPV, and NPV for each mRDT in both the crude form, and stratified by sex, age group, and transmission level. Trends in lower accuracy can be seen for most NPV estimates, sensitivity of the Ragipen test, sensitivity in very low transmission regions, and specificity in high transmission regions.

**Alt Text Figure 1** = Point estimate and whisker plot of 95% Confidence internals for the odds of agreement between the research mRDT (Rapidgen) and standard mRDTs used stratified by transmission intensity. Large confidence intervals denote no statistical difference, but trend is for lower agreement in high transmission settings.

## Data Availability

All data produced in the present study are available upon reasonable request to the authors.

## Acknowledgments

The authors wish to thank the participants and parents/guardians of all the children who participated in the surveys. We acknowledge the contributions of the following project staff and other colleagues who participated in the data collection and/or laboratory processing of the samples: Rule Budodo, Raymond Kitengeso, Rmadhan Moshi, Ruth Mbwambo, Doris Mbata, Salehe Mandai, Gervase A. Chacha, Angelina J. Kisambale, Ezekiel Malecela, Muhidin Kassim, Athanas Mhina, August Nyaki, Juma Tupa, Anangisye Malabeja, Emmanuel Kessy, George Gesase, Tumaini Kamna, Grace Kanyankole, Oswald Oscar, Richard Makono, Ildephonce Mathias, Godbless Msaki, Rashid Mtumba, Gasper Lugela, Gineson Nkya, Daniel Challe, Richard Malisa, Sawaya Msangi, Ally Idrisa, Francis Chambo, Kusa Mchaina, Neema Barua, Christian Msokame, Rogers Msangi, Salome Simba, Hatibu Athumani, Mwanaidi Mtui, Rehema Mtibusa, Juma Akida, Ambele Lyatinga and Tilaus Gustav. The finance, administrative and logistic support team at NIMR: Christopher Masaka, Millen Meena, Beatrice Mwampeta, Gracia Sanga, Neema Manumbu, Halfan Mwanga, Arison Ekoni, Twalipo Mponzi, Pendael Nasary, Denis Byakuzana, Alfred Sezary, Emmanuel Mnzava, John Samwel, Daud Mjema, Seth Nguhu, Thomas Semdoe, Sadiki Yusuph, Alex Mwakibinga, Rodrick Ulomi and Andrea Kimboi. Management of the National Institute for Medical Research, National Malaria Control Program and President’s Office-Regional Administration and Local Government (including the regional administrative secretaries of the 10 regions and district officials) and staff from all 100 health facilities. Technical and logistics support from the Bill and Melinda Gates Foundation team is highly appreciated.

## Disclosures regarding real or perceived conflicts of interest

JBP reports research support from Gilead Sciences, non-financial support from Abbott Laboratories, and consulting for Zymeron Corporation, all outside the scope of the manuscript. All other authors declare no competing interests. Generative AI was used in the writing of this manuscript. The authors take full responsibility for the content.

