## Supplemental Tables for "Varying Malaria Rapid Diagnostic Test Accuracy by Regional Transmission Level and Demographics in Tanzania"

| Supplemental Table 1. Test Accuracy of mRDTs compared to qPCR (qPCR parasitemia cutoff below 50) | | | | |
| --- | --- | --- | --- | --- |
| *Crude Analyses* | |  |  |  |
| mRDT Test Type | Sensitivity | Specificity | Positive Predictive Value (PPV)* | Negative Predictive Value (NPV)* |
| Care Start | 0.874 | 0.888 | 0.935 | 0.793 |
| First Response | 0.830 | 0.876 | 0.877 | 0.892 |
| SD Bioline | 0.871 | 0.897 | 0.940 | 0.791 |
| Rapigen | 0.735 | 0.973 | 0.969 | 0.765 |
| *Stratified by Biological Sex* | |  |  |  |
| Male |  |  |  |  |
| Care Start | 0.869 | 0.900 | 0.935 | 0.806 |
| First Response | 0.832 | 0.871 | 0.874 | 0.827 |
| SD Bioline | 0.892 | 0.865 | 0.934 | 0.790 |
| Rapigen | 0.760 | 0.965 | 0.962 | 0.775 |
| Female |  |  |  |  |
| Care Start | 0.878 | 0.878 | 0.945 | 0.783 |
| First Response | 0.828 | 0.881 | 0.879 | 0.830 |
| SD Bioline | 0.848 | 0.922 | 0.946 | 0.791 |
| Rapigen | 0.714 | 0.981 | 0.976 | 0.757 |
| *Stratified by Age Group* | |  |  |  |
| Children (<5) |  |  |  |  |
| Care Start | 0.928 | 0.810 | 0.906 | 0.850 |
| First Response | 0.848 | 0.834 | 0.860 | 0.820 |
| SD Bioline | 0.867 | 0.873 | 0.931 | 0.767 |
| Rapigen | 0.697 | 0.972 | 0.968 | 0.730 |
| School Aged (5-16) ^†^ |  |  |  |  |
| Care Start | 0.864 | 0.886 | 0.946 | 0.738 |
| First Response | 0.871 | 0.845 | 0.912 | 0.781 |
| SD Bioline | 0.882 | 0.764 | 0.920 | 0.677 |
| Rapigen | 0.816 | 0.952 | 0.971 | 0.727 |
| Adult (>16) |  |  |  |  |
| Care Start | 0.829 | 0.946 | 0.958 | 0.791 |
| First Response | 0.787 | 0.914 | 0.878 | 0.845 |
| SD Bioline | 0.865 | 0.951 | 0.960 | 0.837 |
| Rapigen | 0.723 | 0.979 | 0.969 | 0.800 |
| *For these analyses, the prevalence of the usage of mRDT tests compared to qPCR in the sample population is inherent to the calculation. For a population with differing levels of mRDT test administration, the PPV and NPV will be different. | | | | |
| ^†^ Based on the age range used in the 2017 Tanzania National School Children Survey | | | | |

Supplemental Table 2. mRDT Accuracy v. qPCR, Stratified by Malaria Transmission Levels in Tanzania 2021 with qPCR Parasitemia Threshold

| mRDT Test Type | Sensitivity | Specificity | Positive Predictive Value (PPV)* | Negative Predictive Value (NPV)* |
| --- | --- | --- | --- | --- |
| Very Low |  |  |  |  |
| Care Start^†^ | 0.783 | 0.986 | 0.947 | 0.932 |
| First Response | 0.635 | 0.916 | 0.777 | 0.844 |
| SD Bioline | 0.875 | 0.931 | 0.933 | 0.872 |
| Rapigen | 0.658 | 0.987 | 0.968 | 0.833 |
| Low |  |  |  |  |
| Care Start^†^ | 0.932 | 0.885 | 0.965 | 0.793 |
| First Response | 0.893 | 0.826 | 0.893 | 0.826 |
| SD Bioline | 0.871 | 0.897 | 0.951 | 0.753 |
| Rapigen | 0.815 | 0.962 | 0.973 | 0.754 |
| Moderate |  |  |  |  |
| Care Start^†^ | 0.905 | 0.600 | 0.915 | 0.571 |
| First Response | 0.931 | 0.863 | 0.959 | 0.786 |
| SD Bioline | 0.840 | 0.869 | 0.940 | 0.688 |
| Rapigen | 0.822 | 0.900 | 0.962 | 0.624 |
| High |  |  |  |  |
| Care Start^†^ | 0.725 | 0.833 | 0.906 | 0.577 |
| First Response^†^ | 0.904 | 0.764 | 0.894 | 0.783 |
| SD Bioline | 0.906 | 0.740 | 0.932 | 0.667 |
| Rapigen | 0.681 | 0.950 | 0.974 | 0.524 |
| *For these analyses, the prevalence of the usage of mRDT tests compared to qPCR in the sample population is inherent to the calculation. For a population with differing levels of mRDT test administration, the PPV and NPV will be different. | | | | |
| ^†^ These strata had small sample sizes; Very Low Care Start (n = 93), Low Care Start (n=114 ), Moderate Care Start (n = 115), High Care Start (n = 58), High SD Bioline (n = 133) | | | | |
